# Immunogenicity of Oxford-AstraZeneca COVID-19 vaccine in Vietnamese healthcare workers

**DOI:** 10.1101/2021.07.08.21260162

**Authors:** Nguyen Van Vinh Chau, Lam Anh Nguyet, Nguyen Thanh Truong, Le Mau Toan, Nguyen Thanh Dung, Le Manh Hung, Mai Thanh Nhan, Dinh Nguyen Huy Man, Nghiem My Ngoc, Huynh Phuong Thao, Tran Nguyen Hoang Tu, Huynh Kim Mai, Do Thai Hung, Nguyen Thi Han Ny, Le Kim Thanh, Nguyen To Anh, Nguyen Thi Thu Hong, Le Nguyen Truc Nhu, Lam Minh Yen, Marc Choisy, Tran Tan Thanh, Guy Thwaites, Le Van Tan, OUCRU COVID-19 Research Group

**Author notes:** Correspondence: Nguyen Van Vinh Chau, Le Van Tan. Members of the groups are listed in the acknowledgments (OUCRU COVID-19 Research Group).

## Abstract

We studied the immunogenicity of Oxford-AstraZeneca vaccine in Vietnamese healthcare workers. We collected blood samples before each dose, at 14 days after each dose, and month 1 and 3 after dose 1 from each participant alongside demographics data. We measured neutralizing antibodies using a surrogate virus neutralization assay. The 554 study participants (136 males and 418 females) were aged between 22-71 years (median: 36 years). 104 and 94 out of 144 selected participants were successfully followed up at 14 days after dose 2 and 3 months after dose 1, respectively. Neutralizing antibodies increased after each dose, with the sero-conversion rate reaching 98.1% (102/104) at 14 days after dose 2. At month 3 after dose 1, neutralizing antibody levels decreased, while 94.7% (89/94) of the study participants remained seropositive. Oxford-AstraZeneca COVID-19 vaccine is immunogenic in Vietnamese healthcare workers. The requirement for a third dose warrants further research.

## BACKGROUND

Severe acute respiratory syndrome coronavirus 2 (SARS-CoV-2) is the cause of the ongoing COVID-19 pandemic [1]. Since its first detection in Wuhan, China in late 2019, SARS-CoV-2 has now become an endemic virus globally. Vaccine is thus the most plausible approach to return to the pre-pandemic life. As such, vaccine development has ramped up globally over the last year. As of 1^st^ June 2021, a total of 185 and 102 vaccine candidates are under the pre-clinical and clinical development phases, respectively [2]. Additionally, seven vaccines have received the WHO approval for emergency use [2]. Approved vaccines have been rapidly deployed globally. And as of 26^th^ July 2021, over 3.8 billion doses of COVID-19 vaccines have been administered worldwide. Vietnam received the first doses of Oxford-AstraZeneca vaccine in early March 2021. As of 18^st^ July 2021 over four million doses have been administered in Vietnam; the majority was Oxford-AstraZeneca vaccines [3].

Although a vaccine must fulfill the required efficacy criteria in order to receive an approval for use in humans, the rapid development and deployment of COVID-19 vaccines worldwide necessitate follow up studies to better understand the development and persistence of vaccine-induced immunity in different populations. Such knowledge is critical to inform the global vaccination strategies and the development of next-generation vaccines.

Despite the current surge, which has been escalating since the second week of May 2021, Vietnamese people remained relatively naïve to SARS-CoV-2 infections [4, 5]. As of 18^th^ July 2021, a total of 31,391 PCR confirmed cases have been reported in Vietnam, a country of over 97 million people [3]. Therefore Vietnam is an ideal setting for vaccine evaluation study as the results naturally reflect the immunity induced by COVID-19 vaccines. There has been no report about the immunogenicity of the Oxford-AstraZeneca COVID-19 vaccine from Southeast Asia. We studied the immunogenicity of Oxford-AstraZeneca COVID-19 vaccine in a cohort of 554 healthcare workers of an infectious diseases hospital in southern Vietnam.

## METHODS

### Setting and COVID-19 vaccine rollout in Vietnam

The present study was conducted at the Hospital for Tropical Diseases (HTD) in HCMC. HTD is a 550-bed tertiary referral hospital for patients with infectious diseases (including COVID-19) in southern Vietnam [6].

Vietnam received the first 117,000 doses of Oxford-AstraZeneca COVID-19 vaccine in early March 2021. The window time between two doses was set for 4 weeks, with some variation depending on the availability of the vaccine. According to the Vietnamese Ministry of Health, high-risk groups, especially frontline healthcare workers, were prioritized for vaccination (Supplementary Materials). HTD members of staff were eligible for vaccination and were the first in Vietnam to receive a COVID-19 vaccine in March 2021.

### Data collection

We collected demographics and 3ml of blood from the study participants. Blood sampling was scheduled for 7 time points, including before each dose, 14 days after each dose, and month 1, 3, 6 and 12 after vaccination. After day 28 of the first dose, blood sampling was narrowed down to a subgroup of 144 individuals randomly selected from the study participants for subsequent follow up. The present report focused on the period from baseline to month 3 after the first dose.

### Neutralizing antibody measurement

Neutralizing antibodies were measured using an FDA EUA approved assay, namely SARS-CoV-2 Surrogate Virus Neutralization Test (sVNT) (GenScript, USA). Prior to testing, plasma samples were first diluted 1:10 and then inactivated at 56°C for 30 minutes. The experiments were carried out according the manufacturer’s instruction. The obtained results were expressed as percentage of inhibition with the 30% cut-off applied. The percentage of inhibition measured by sVNT has been shown to well correlate with the neutralizing antibody tiers measured by the conventional plaque reduction neutralization assay [7].

### Neutralizing antibody data from cases of natural infection

To compare the development of neutralizing antibodies induced by vaccination against that of natural infection, we included data from 11 Vietnamese patients who had mild or asymptomatic infections. Details about these individuals and neutralizing antibody measurement were detailed in our recent report [8].

### Statistical analysis

We used Fisher exact, χ^2^ or Mann-Whitney U test to compare between groups (when appropriate). Logistic regression was used to assess association between the probability of having detectable neutralizing antibodies and age. Linear regression was used to assess the association between neutralizing antibodies levels and age. The analyses were carried using Prism 9.0.2 (graphpad.com).

### Ethics

The study was approved by the Institutional Review Board of HTD and the Oxford Tropical Research Ethics Committee, University of Oxford, UK. Written informed consents were obtained all the participants.

## RESULTS

### Demographics of the study participants

A total 649/894 (72.6%) HTD staff consented to participate in the vaccine evaluation study. 554/649 (85.4%) participants were successfully followed-up up to day 28 after the first dose and were thus included for analysis as whole group. The 554 study participants were aged between 22 and 71 years (median: 37 years). Females were predominant, accounting for 75.4% (418/554) (Table 1)

Of the 144 participants of the subgroup, 104 (72.2%) and 94 (65.3%) were successfully followed-up up to 14 days after the second dose and 3 months after the first dose, respectively. The age and gender distributions of these subgroups were comparable with that of the whole group (Table 1). The window time between the first and the second dose was six weeks.

**Table 1:** Demographics of the study participants

|  | **Whole group (N=554)** | **Subgroup (N=104)** | **Subgroup (N=94)** |
| --- | --- | --- | --- |
| Male, n (%) | 136 (25) | 25 (24) | 21 (22) |
| Female, n (%) | 418 (75) | 79 (76) | 73 (78) |
| Median age in years (range) | 36 (22 – 71) | 37 (24 – 65) | 37 (24-65) |
| Age groups |  |  |  |
| 20-39, n (%) | 332 (60) | 57 (55) | 50 (53) |
| 40-60, n (%) | 217 (39) | 46 (44) | 43 (46) |
| 61-71, n (%) | 5 (1) | 1 (1) | 1 (1) |

### Development of detectable neutralizing antibodies

Because HTD members of staff were naïve to SARS-CoV-2 infection [4], we first focused our neutralizing antibody measurement on the baseline samples collected before the first dose of the subgroup. Indeed, at baseline, none of the 104 study participants had detectable neutralizing antibodies (Table 2). At day 14 and 28 after the first dose, the proportions of the study participants with detectable neutralizing antibodies increased from 27.3% (151/554) to 78.0% (432/554), respectively among all 554 individuals of the whole group. The proportion of the study participants with detectable neutralizing antibodies reached 98.1% (102/104) at 14 days after the second dose, and then slightly dropped to 94.7% (89/94) at month 3 after the first dose (i.e. six weeks after the second dose) (Table 2).

**Table 2:** The proportion of study participants with detectable neutralizing antibodies after vaccination

| Time point | Whole group |  |  |  | Subgroup |  |  |  |
| --- | --- | --- | --- | --- | --- | --- | --- | --- |
|  | Total<br>(N=554) | Male<br>(N=136) | Female<br>(N=418) | P value* | Total<br>(N=104) | Male<br>(N=25) | Female<br>(N=79) | P value* |
| Baseline, n (%) | 0 | 0 | 0 | NA | 0 | 0 | 0 | NA |
| 14 days after dose 1, n (%) | 151 (27.3) | 40 (29.4) | 111 (26.6) | 0.52 | 31 (29.8) | 10 (40.0) | 21 (26.6) | 0.20 |
| 28 days after dose 1, n (%) | 432 (78.0) | 97 (71.3) | 335 (80.1) | 0.031 | 82 (78.8) | 20 (80.0) | 62 (78.5) | 0.87 |
| Before dose 2, n (%) | N/A | N/A | N/A | N/A | 73 (70.2) | 17 (68.0) | 56 (70.1) | 0.78 |
| 14 days after dose 2, n (%) | N/A | N/A | N/A | N/A | 102 (98.1) | 24 (96.0) | 78 (98.7) | 0.43 |
| Month 3 after the first dose** | NA | NA | NA | NA | 89 (94.7) | 21 (95.5) | 68 (94.4) | 1 |

**Notes to Table 2:** \*for comparison between males and females, NA: non-applicable, \*\*n=94 (male: 22 and females: 72)

### Kinetics of neutralizing antibody levels

After the first dose, neutralizing antibody levels measured at day 28 were significantly higher than that measured at day 14 (Figure 1A), but comparable with that measured at week 6 (Figure 1B). At day 14 after the second dose, neutralizing antibodies significantly increased, and were comparable with that obtained from Vietnamese people with asymptomatic or mild infection (Figure 1B). At month 3 after the first does neutralizing antibody levels were significantly lower than that measured at 14 days after the second dose (Figure 1B)

**Figure 1:**
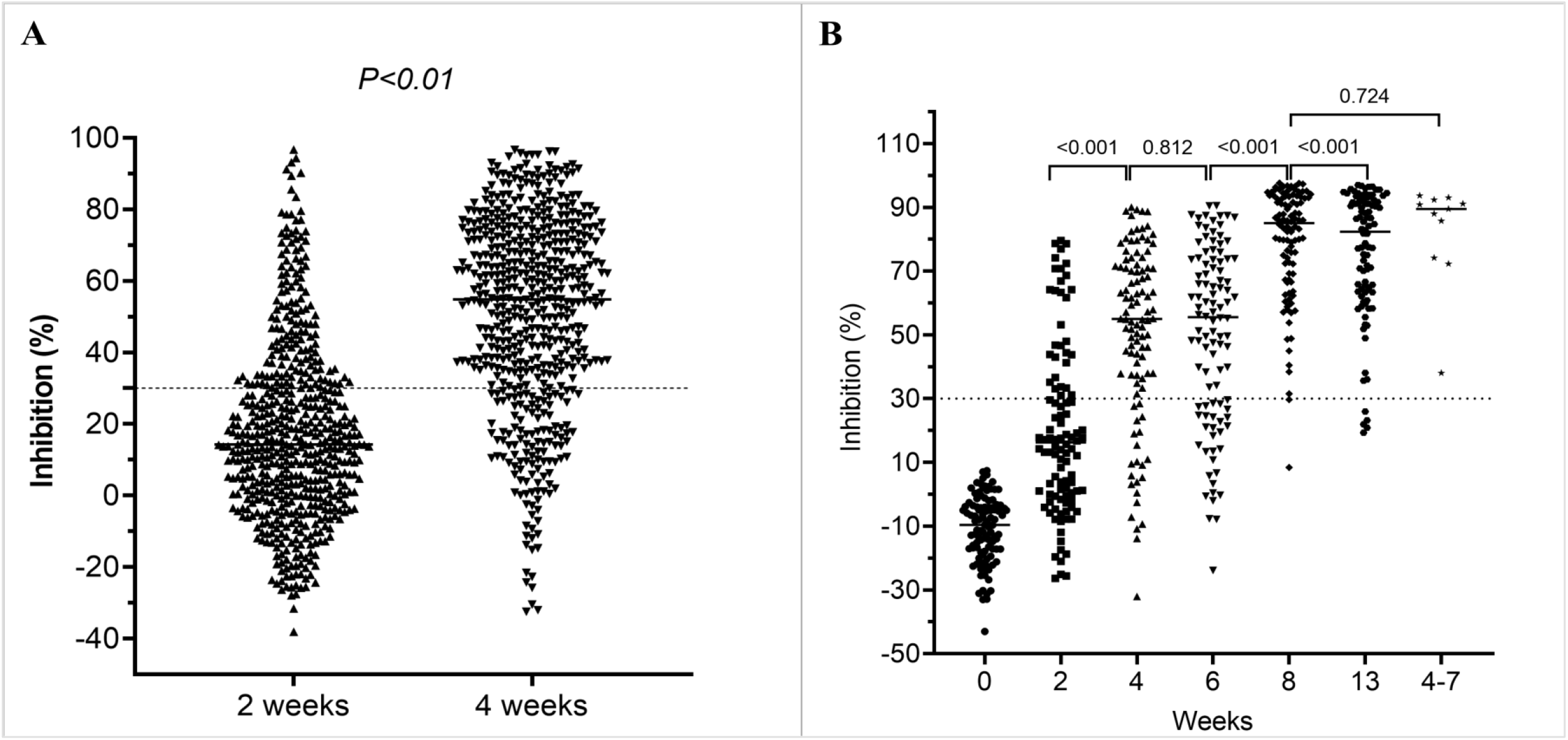
Development of neutralizing antibodies levels after vaccination, **A**) Neutralizing antibody levels measured at 14 and 28 days after the first dose of 554 study participants; Neutralizing antibody levels measured at time points from baseline to month 3 after the firs dose of the subgroups; **Notes to figure 1B**: Data on neutralizing antibody levels obtained from 11 convalescent sera collected at week 4-7 (last column) from cases with mild or asymptomatic infection was included as references.

### Neutralizing antibodies vs. age and gender

At day 14 after the first dose, the development and levels of detectable neutralizing antibodies among 554 study participants were negatively correlated with age. This difference was less profound at day 28 after dose 1, especially with regard to the development of detectable neutralizing antibodies (Figure 2). At these corresponding time points, similar trends were also observed among individuals of the subgroup, but the difference was not significant (Figure 3 and Supplementary Figure 1), likely because of the small sample size. At 14 days after the second dose and month 3 after the first dose, the proportion of individuals with detectable neutralizing antibodies was similar across age groups (Supplementary Figure 1B&C).

**Figure 2:**
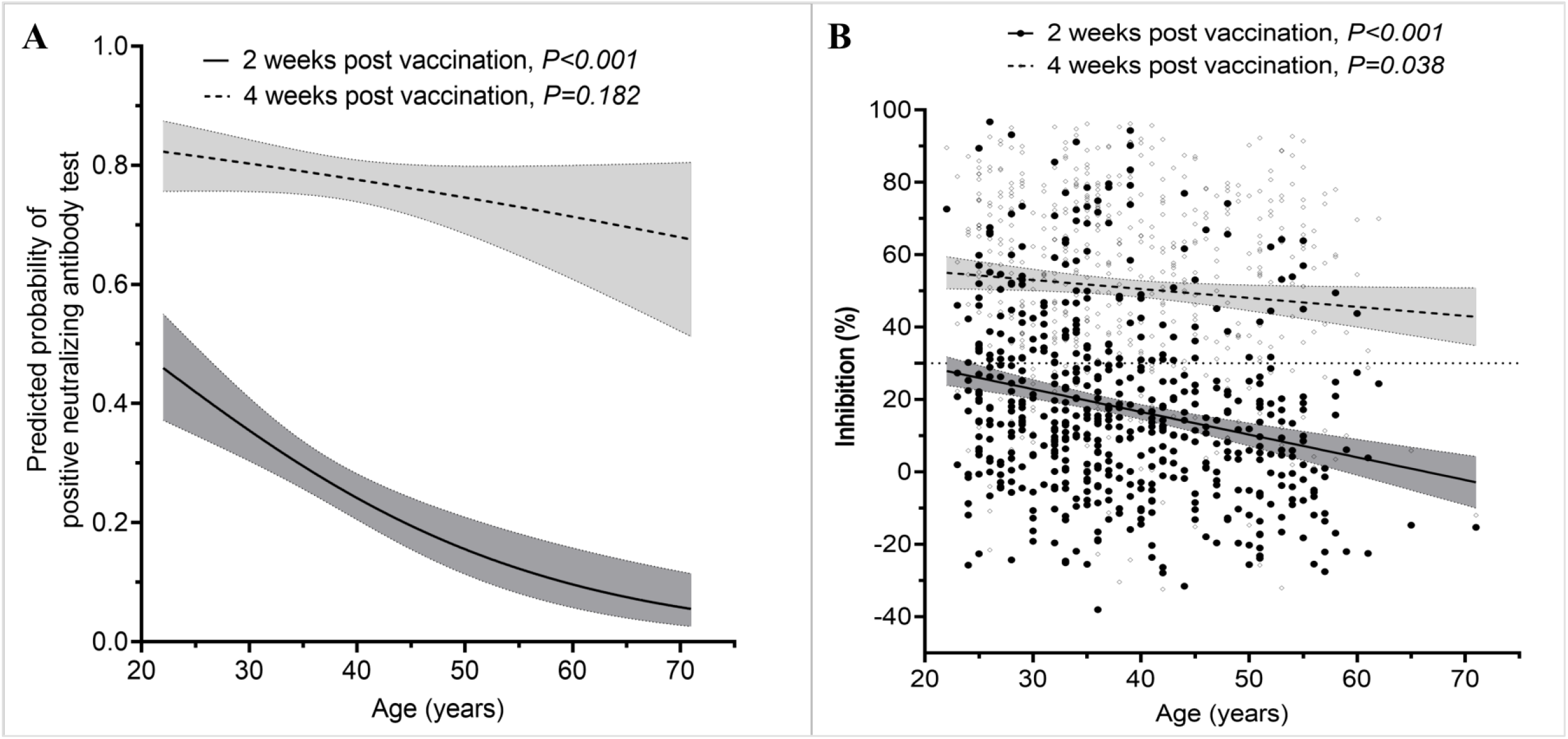
The associations between neutralizing antibody level and age, **A**) Association between age and the probabilities of having detectable neutralizing antibodies at 14 and 28 days after the first does of 554 study participants; **B)** Association between age and neutralizing antibody levels measured at 14 and 28 days after the first dose of 554 study participants **Notes to Figure 2**: Black circles: data for 2 week time point and grey circles: data for 4 week time point. Shaded areas indicate 95% of confident intervals.

**Figure 3:**
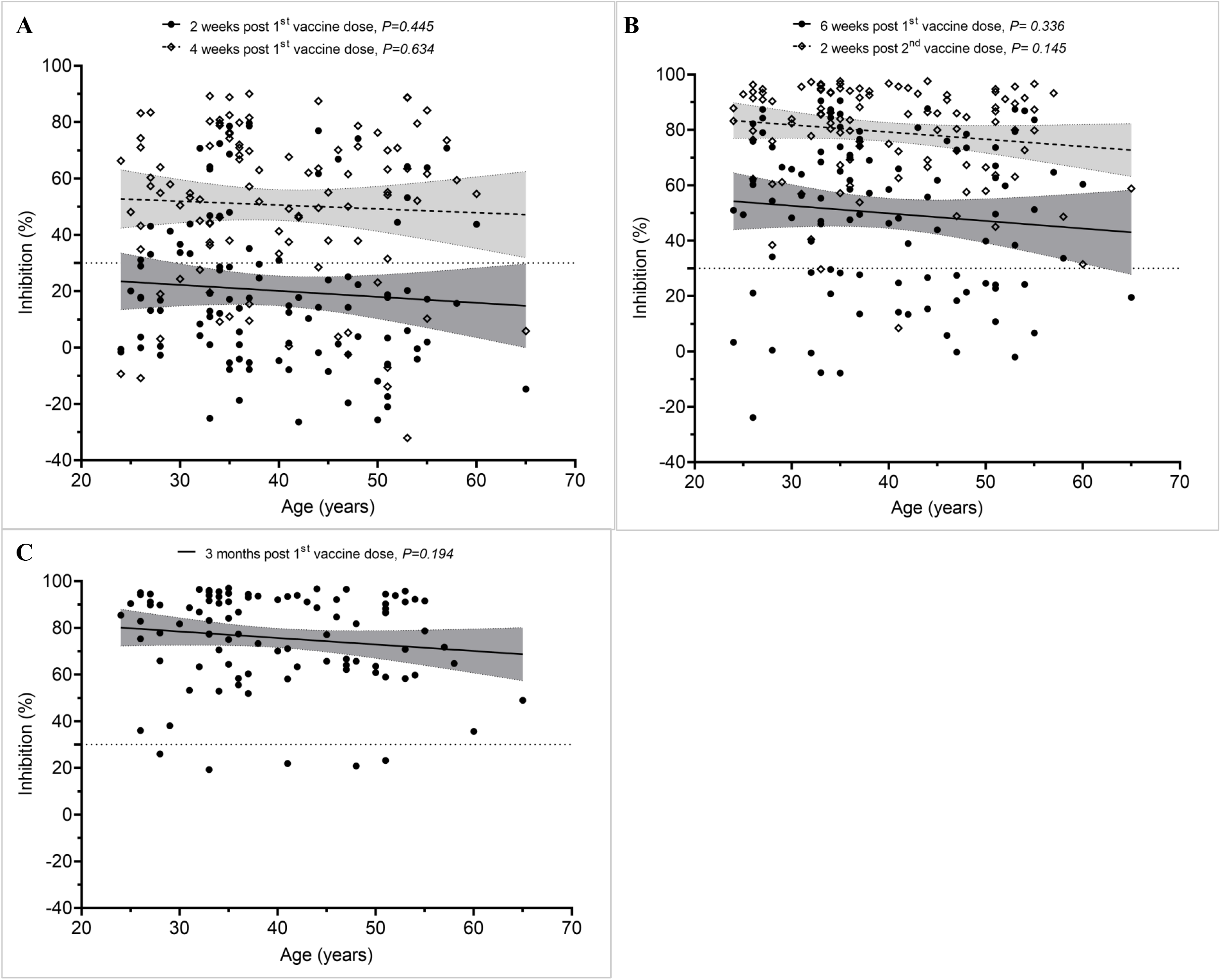
Neutralizing antibody levels of participants selected for assessment of the impact of the second dose. **A**) At 2 and 4 weeks after the first dose (n=104), **B**) Before the second dose (i.e.6 weeks after the first dose) and 2 weeks after the second dose (n=104) and **C**) at month 3 after the first dose (n=94). **Notes to Figure 3**: Shaded areas indicate 95% of confident intervals.

In terms of gender, with the exception of day 28 after the first dose, neutralizing antibody levels and the proportion of study participants with detectable neutralizing antibodies were comparable between males and females (Table 2 and Supplementary Figure 2).

## DISCUSSION

We report the immunogenicity of Oxford-AstraZeneca COVID-19 vaccine in a cohort of 554 Vietnamese healthcare workers who were naïve to SARS-CoV-2 infection. We showed that Oxford-AstraZeneca COVID-19 vaccine is immunogenic in Vietnamese people. Neutralizing antibodies increased after each dose, with the sero-conversion rate reaching 98.1% (102/104) at 14 days after dose 2. At month 3 after dose 1, neutralizing antibody levels decreased, while 94.7% (89/94) of the study participants remained seropositive.

Findings from the original phase 2/3 trial showed that spike protein specific IgG developed within two weeks after vaccination, and at 14 days after the second dose its titers increased with a sero-conversion rate of 208/209 (>99%) [9]. Consistently, our study showed the development and the levels of neutralizing antibodies significantly increases after each dose, with the former reaching 98.1% at 14 days after the second dose. Parallel with these reports are real-word data from the UK showing that the administration of the second dose increased protection against SARS-CoV-2 infection from 65% by dose 1 to 70% by dose 2 among vaccine recipients [10]. A single dose of Oxford-AstraZeneca or Pfizer COVID-19 vaccines reduced COVID-19 hospital admissions among vaccine recipients by 88% and 91%, respectively in Scotland [11].

Older individuals, especially those 80 years or above, without prior infection had lower levels of neutralizing antibodies induced by the first dose than younger adults [12, 13]. These age-dependent responses were most profound within the first 3 weeks after vaccination, but were resolved by the administration of the second dose [12]. Although similar trends were observed in our study, at day 28 after the first dose, the differences in our study were negligible, especially in terms of the sero-conversion rate. None of our study participant was older than 71 years, explaining why the observed differences were less profound as compared to the UK population based study.

The results provide reassuring evidence for the effectiveness of the proposed vaccination strategy aiming at prioritizing the first dose for as many people as possible in the first instance [14]. However, the data also emphasize the importance of the second dose [15], especially in older people, while it remains unknown whether the third dose is needed to provide long-term protection. A decline in antibody titers was recorded at week 8-12 after the first two doses among 75 study participants in the UK [16]. But the administration of the third dose helped boost the immune response. Antibody waning is presumably more profound among individuals without prior infection. Follow study is therefore critical to assess the levels of antibody waning among our study participants and correlates of protection, especially in the context of the rapid spread of the Delta variant globally.

The strength of our study includes that it was conducted in a naïve population, with no prior SARS-CoV-2 infections [4]. Thus our data more naturally reflect the immunity profiles induced by Oxford-AstraZeneca COVID-19 vaccine. Additionally, although the correlates of protection for COVID-19 vaccine remain to be determined, neutralizing antibodies are considered to be the most reliable surrogates [17]. Therefore by measuring neutralizing antibodies, our findings more accurately reflect the potential of correlates of protection.

Our study has some limitations. First, we did not study cellular immunity, especially T cell response. Cellular immunity has been increasingly recognized to play a role in the pathogenicity and immune response of SARS-CoV-2 infection [18]. Therefore, the durability of both cellular and humoral immune responses should be further explored. Second, due to the age and gender structure in nature of HTD staff, we did not include participants older than 71 years and females were predominant among our study subjects. Of note, compared with males, females seemed to better respond to Oxford-AstraZeneca COVID-19 vaccine at day 28 after the first dose, which merits further research In summary, Oxford-AstraZeneca COVID-19 vaccine is immunogenic in Vietnamese healthcare workers who were naïve to SARS-CoV-2 infection. Neutralizing antibody levels decreased at month 3 after vaccination. The requirement for a third dose warrants further research. These data are critical to informing the deployment of COVID-19 vaccine in Vietnam and beyond.

## Data Availability

The authors confirm that the data supporting the findings of this study are available within the article [and/or its supplementary materials].

## ACKNOWLEDGEMENTS

This study was funded by the Wellcome Trust of Great Britain (106680/B/14/Z and 204904/Z/16/Z) and NIH/NIAID (HHSN272201400007C, Subcontract No. S000596-JHU)..We thank our colleagues at the Hospital for Tropical Diseases in Ho Chi Minh City, Vietnam for their participations in this study.

## OUCRU COVID-19 Research Group

### Hospital for Tropical Diseases, Ho Chi Minh City, Vietnam

Nguyen Van Vinh Chau, Nguyen Thanh Dung, Le Manh Hung, Huynh Thi Loan, Nguyen Thanh Truong, Nguyen Thanh Phong, Dinh Nguyen Huy Man, Nguyen Van Hao, Duong Bich Thuy, Nghiem My Ngoc, Nguyen Phu Huong Lan, Pham Thi Ngoc Thoa, Tran Nguyen Phuong Thao, Tran Thi Lan Phuong, Le Thi Tam Uyen, Tran Thi Thanh Tam, Bui Thi Ton That, Huynh Kim Nhung, Ngo Tan Tai, Tran Nguyen Hoang Tu, Vo Trong Vuong, Dinh Thi Bich Ty, Le Thi Dung, Thai Lam Uyen, Nguyen Thi My Tien, Ho Thi Thu Thao, Nguyen Ngoc Thao, Huynh Ngoc Thien Vuong, Huynh Trung Trieu Pham Ngoc Phuong Thao, Phan Minh Phuong

### Oxford University Clinical Research Unit, Ho Chi Minh City, Vietnam

Dong Thi Hoai Tam, Evelyne Kestelyn, Donovan Joseph, Ronald Geskus, Guy Thwaites, Ho Quang Chanh, H. Rogier van Doorn, Ho Van Hien, Ho Thi Bich Hai, Huynh Le Anh Huy, Huynh Ngan Ha, Huynh Xuan Yen, Jennifer Van Nuil, Jeremy Day, Katrina Lawson, Lam Anh Nguyet, Lam Minh Yen, Le Dinh Van Khoa, Le Nguyen Truc Nhu, Le Thanh Hoang Nhat, Le Van Tan, Sonia Lewycka Odette, Louise Thwaites, Marc Choisy, Mary Chambers, Ngo Thi Hoa, Nguyen Thanh Thuy Nhien, Nguyen Thi Han Ny, Nguyen Thi Kim Tuyen, Nguyen Thi Phuong Dung, Nguyen Thi Thu Hong, Nguyen Xuan Truong, Phan Nguyen Quoc Khanh, Phung Le Kim Yen, Phung Tran Huy Nhat, Sophie Yacoub, Thomas Kesteman, Nguyen Thuy Thuong Thuong, Tran Tan Thanh, Tran Tinh Hien, Vu Thi Ty Hang

## Supplementary Materials

### List of groups prioritized for COVID-19 vaccination in Vietnam

1. Frontline healthcare workers of COVID-19, including people whose work is to deal with COVID-19 prevention and control work (members of COVID-19 steering committees at all levels, staff at state-run quarantine sites, people conducting contract tracing and epidemiological investigations, volunteers, reporters among others), military and public security forces
2. Vietnamese diplomats, customs and immigration officers
3. Essential service workers in sectors such as aviation, transport, tourism, electricity and water supply
4. Teachers and individuals working at education and training facilities, and those working at State agencies with regular contact with various people
5. People with chronic diseases or aging above 65
6. Residents in outbreak hotspots in Vietnam
7. Poor people, Policy beneficiaries
8. Those who will be sent abroad for learning and working
9. Other people determined by the Ministry of Health

**Supplementary Figure 1:**
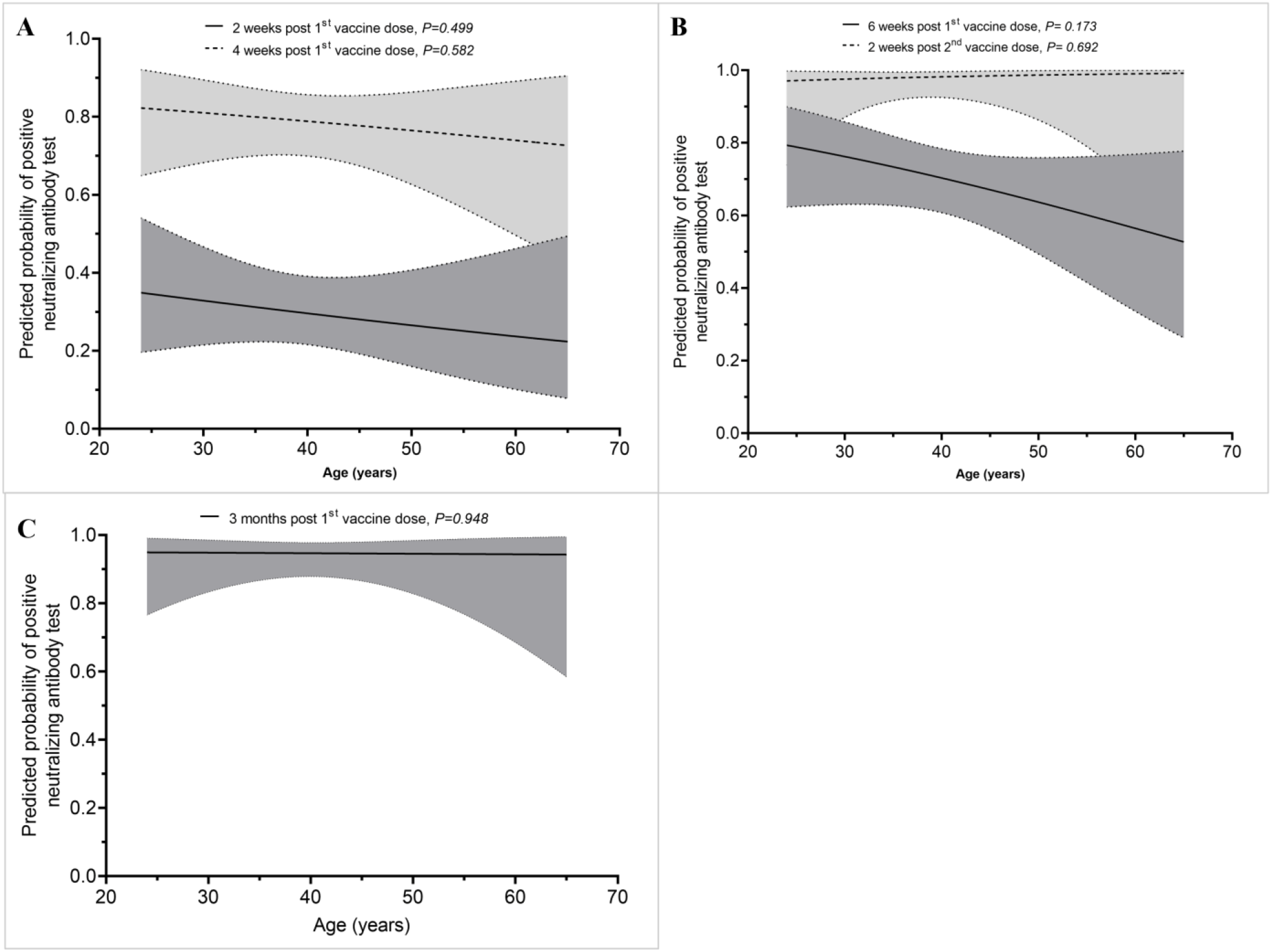
Probability of having detectable neutralizing antibodies among the study participants selected for assessment of the impact of the second dose. **A**) At 2 and 4 weeks after the first dose (n=104), **B**) Before the second dose (i.e.6 weeks after the first dose) and 2 weeks after the second dose (n=104) and **C**) at month 3 after the first dose (n=94). Shaded areas indicate 95% of confident intervals.

**Supplementary Figure 2:**
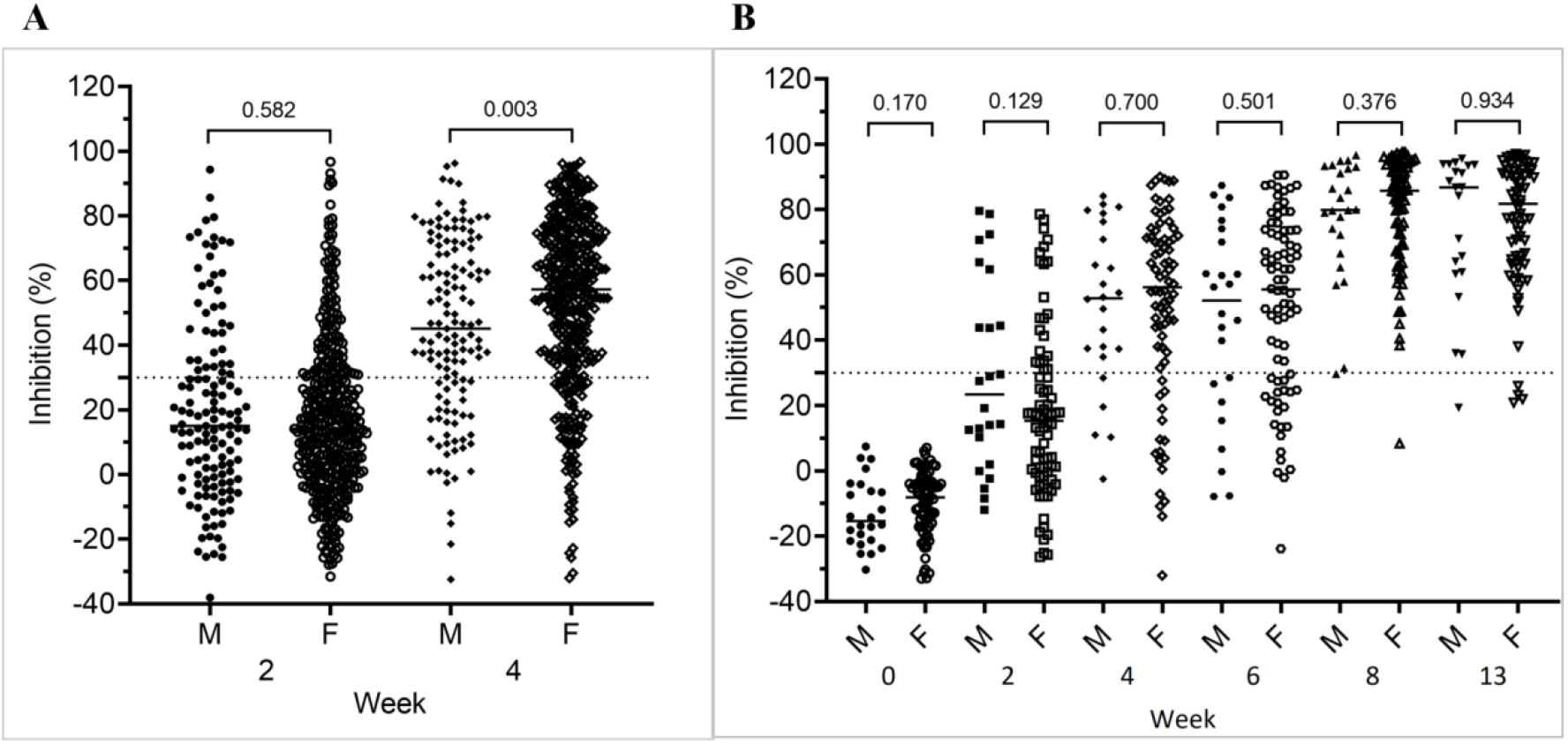
Association between neutralizing antibody levels and gender. **A**) At 2 and 4 weeks after the first dose of the whole group (n=554), **B**) From baseline to month three after the first dose of the subgroup

